# Using Machine Learning of Clinical Data to Diagnose COVID-19

**DOI:** 10.1101/2020.06.24.20138859

**Authors:** Wei Tse Li, Jiayan Ma, Neil Shende, Grant Castaneda, Jaideep Chakladar, Joseph C. Tsai, Lauren Apostol, Christine O. Honda, Jingyue Xu, Lindsay M. Wong, Tianyi Zhang, Abby Lee, Aditi Gnanasekar, Thomas K. Honda, Selena Z. Kuo, Michael Andrew Yu, Eric Y. Chang, Mahadevan “Raj” Rajasekaran, Weg M. Ongkeko

**Author notes:** Authors contributed equally (Wei Tse Li, Jiayan Ma, Neil Shende).

## Abstract

The recent pandemic of Coronavirus Disease 2019 (COVID-19) has placed severe stress on healthcare systems worldwide, which is amplified by the critical shortage of COVID-19 tests. In this study, we propose to generate a more accurate diagnosis model of COVID-19 based on patient symptoms and routine test results by applying machine learning to reanalyzing COVID-19 data from 151 published studies. We aimed to investigate correlations between clinical variables, cluster COVID-19 patients into subtypes, and generate a computational classification model for discriminating between COVID −19 patients and influenza patients based on clinical variables alone. We discovered several novel associations between clinical variables, including correlations between being male and having higher levels of serum lymphocytes and neutrophils. We found that COVID-19 patients could be clustered into subtypes based on serum levels of immune cells, gender, and reported symptoms. Finally, we trained an XGBoost model to achieve a sensitivity of 92.5% and a specificity of 97.9% in discriminating COVID-19 patients from influenza patients. We demonstrated that computational methods trained on large clinical datasets could yield ever more accurate COVID-19 diagnostic models to mitigate the impact of lack of testing. We also presented previously unknown COVID-19 clinical variable correlations and clinical subgroups.

## 1. Introduction

COVID-19 is a severe respiratory illness caused by the virus SARS-Cov-2. The scientific community has focused on this disease with near unprecedented intensity. However, the majority of primary studies published on COVID-19 suffered from small sample sizes[1, 2]. While a few primary research studies reported on dozens or hundreds of cases, many more studies reported on less than 20 patients[3, 4]. Therefore, there is an urgent need to collate all available published data on the clinical characteristics of COVID-19 from different studies to construct a comprehensive dataset for gaining insights into the pathogenesis and clinical characteristics of COVID-19. In this study, we aim to perform a large-scale meta-analysis to synthesize all published studies with COVID-19 patient clinical data, with the goal of uncovering novel correlations between clinical variables in COVID-19 patients. We will then apply machine learning to reanalyze the data and construct a computational model for predicting whether someone has COVID-19 based on their clinical information alone.

We believe that the ability of predicting COVID-19 patients based on clinical variables and using an easily accessible computational model would be extremely useful to address the widespread lack of testing capabilities for COVID-19 worldwide. Because many countries and hospitals are not able to allocate sufficient testing resources, healthcare systems are deprived of one of their most effective tools for containing a pandemic: identification of case hotspots and targeted action towards regions and specific individuals with the disease[5]. The scale of the testing shortage calls for methods for diagnosing COVID-19 that use resources local healthcare facilities currently have. We propose the development of a disease prediction model based on clinical variables and standard clinical laboratory tests.

A number of meta-analyses have been done on COVID-19, but almost none of them comprehensively included data from all published studies. Three different meta-analyses, published in February, March, and April of 2020, included data from 10, 8, and 31 articles, respectively[6-8]. We included 151 articles, comprising 413 patients, in our analysis. To the best of our knowledge, no study has performed a large-scale machine learning analysis on clinical variables to obtain a diagnostic model. We believe that our study will be an important step towards leveraging the full extent of published clinical information on COVID-19 patients to inform diagnosis of COVID-19, instead of relying on general guidelines for symptoms that do not take into account the association between different clinical variables.

## 2. Results

### 2.1 Compilation of patient data and summary of clinical variables

After compiling information from 151 published studies, we present a total of 42 different clinical variables, including 21 categorical and 21 continuous variables, that are reported in more than 1 study. Discrete variables include nominal categorical variables like gender, which is 49.49% (194 patients) male and 50.51% (198 patients) female, and ordinal categorical variables like lymphocytes level, of which 86 patients (48.86%) have low levels, 73 patients (41.47%) have normal levels, and 17 patients (9.65%) have high levels. Continuous variables include age, which has a mean of 38.91 years and variance 21.86 years, and serum neutrophil levels, which has a mean of 6.85 x10^9^ cells/L and a variance of 12.63 x10^9^ cells/L. Certain variables, including all counts of blood cell populations, have both ordinal and continuous components. The continuous component describes the raw count of these populations, while the ordinal component describes whether these counts are within normal range, below normal range, or above normal range. Summary for all data are shown in Table 1.

**Table 1:**
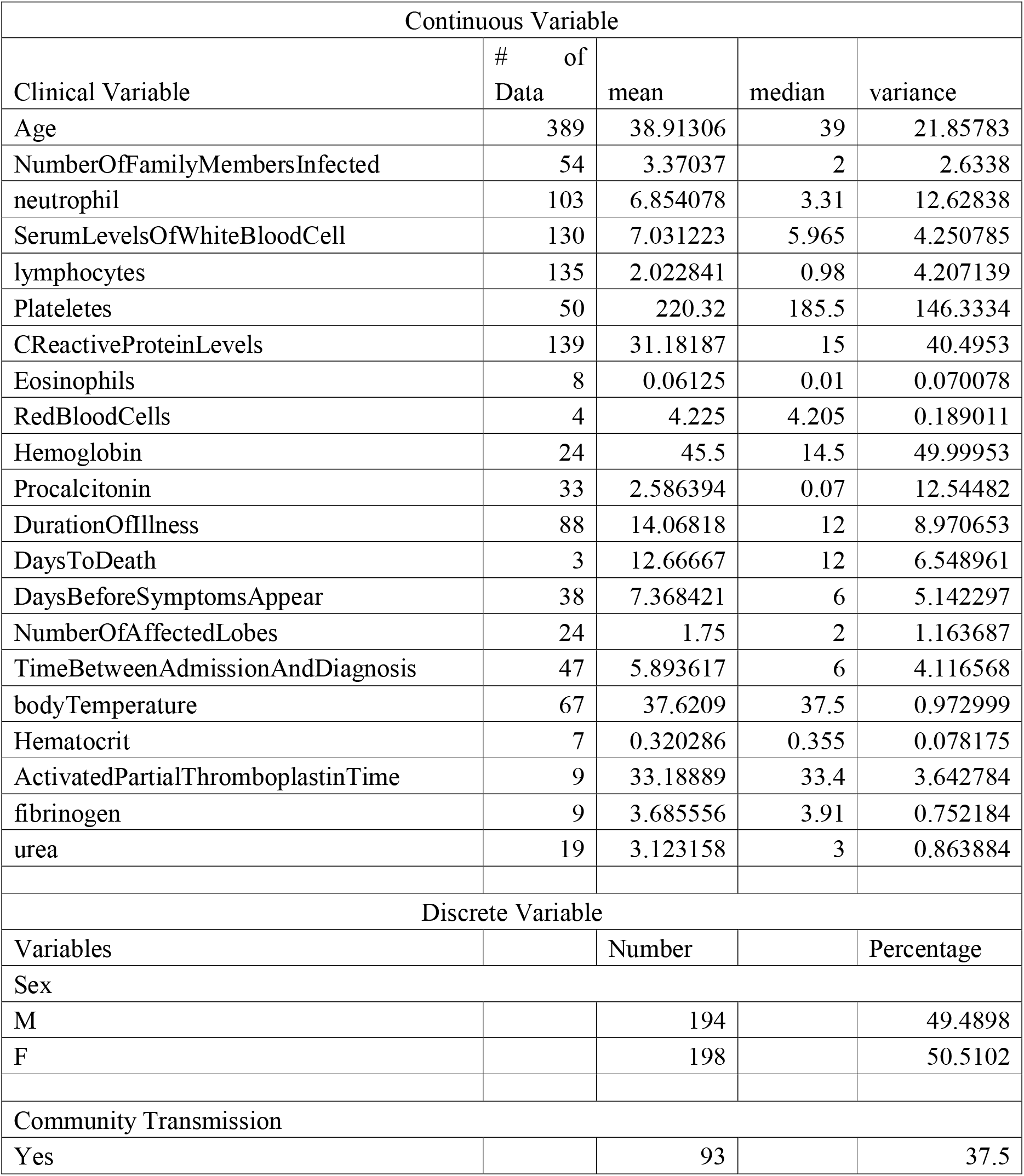
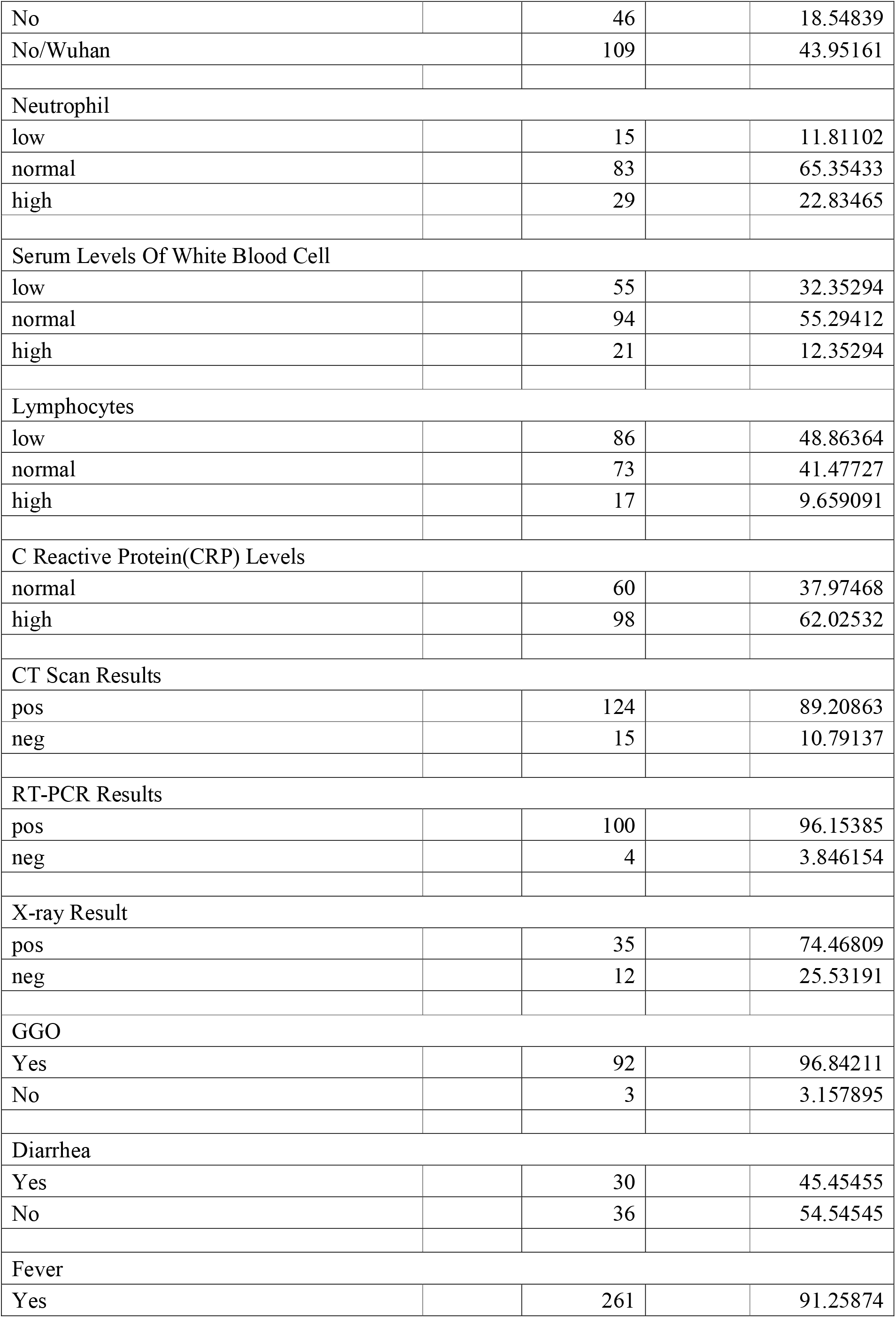
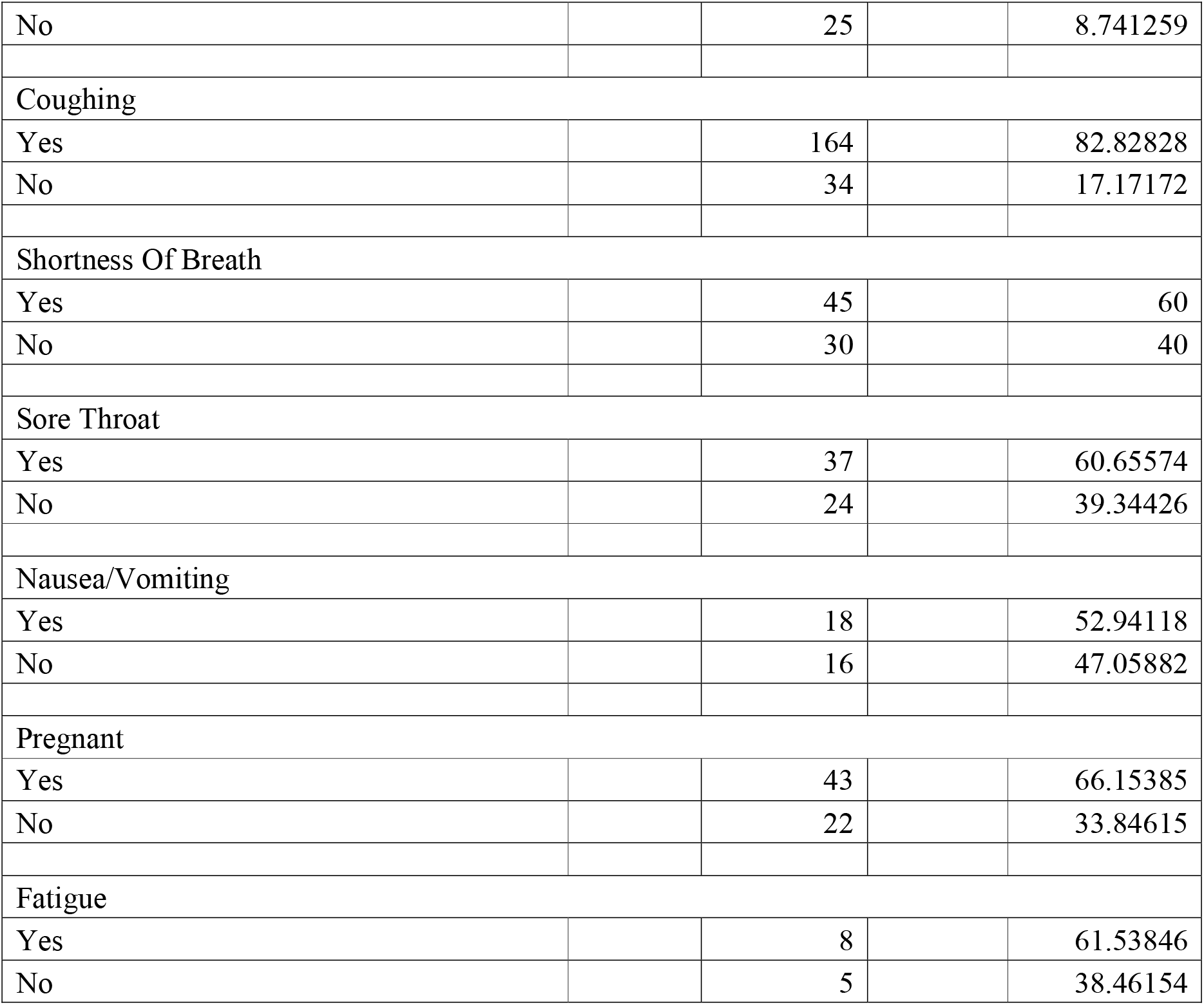
Clinical Variables Summary of Meta-analysis

| Continuous Variable |  |  |  |  |
| --- | --- | --- | --- | --- |
| Clinical Variable | # of Data | mean | median | variance |
| Age | 389 | 38.91306 | 39 | 21.85783 |
| NumberOfFamilyMembersInfected | 54 | 3.37037 | 2 | 2.6338 |
| neutrophil | 103 | 6.854078 | 3.31 | 12.62838 |
| SerumLevelsOfWhiteBloodCell | 130 | 7.031223 | 5.965 | 4.250785 |
| lymphocytes | 135 | 2.022841 | 0.98 | 4.207139 |
| Plateletes | 50 | 220.32 | 185.5 | 146.3334 |
| CReactiveProteinLevels | 139 | 31.18187 | 15 | 40.4953 |
| Eosinophils | 8 | 0.06125 | 0.01 | 0.070078 |
| RedBloodCells | 4 | 4.225 | 4.205 | 0.189011 |
| Hemoglobin | 24 | 45.5 | 14.5 | 49.99953 |
| Procalcitonin | 33 | 2.586394 | 0.07 | 12.54482 |
| DurationOfIllness | 88 | 14.06818 | 12 | 8.970653 |
| DaysToDeath | 3 | 12.66667 | 12 | 6.548961 |
| DaysBeforeSymptomsAppear | 38 | 7.368421 | 6 | 5.142297 |
| NumberOfAffectedLobes | 24 | 1.75 | 2 | 1.163687 |
| TimeBetweenAdmissionAndDiagnosis | 47 | 5.893617 | 6 | 4.116568 |
| bodyTemperature | 67 | 37.6209 | 37.5 | 0.972999 |
| Hematocrit | 7 | 0.320286 | 0.355 | 0.078175 |
| ActivatedPartialThromboplastinTime | 9 | 33.18889 | 33.4 | 3.642784 |
| fibrinogen | 9 | 3.685556 | 3.91 | 0.752184 |
| urea | 19 | 3.123158 | 3 | 0.863884 |
| Discrete Variable |  |  |  |  |
| Variables |  | Number |  | Percentage |
| Sex |  |  |  |  |
| M |  | 194 |  | 49.4898 |
| F |  | 198 |  | 50.5102 |
| Community Transmission |  |  |  |  |
| Yes |  | 93 |  | 37.5 |
| No |  | 46 |  | 18.54839 |
| No/Wuhan |  | 109 |  | 43.95161 |
| Neutrophil |  |  |  |  |
| low |  | 15 |  | 11.81102 |
| normal |  | 83 |  | 65.35433 |
| high |  | 29 |  | 22.83465 |
| Serum Levels Of White Blood Cell |  |  |  |  |
| low |  | 55 |  | 32.35294 |
| normal |  | 94 |  | 55.29412 |
| high |  | 21 |  | 12.35294 |
| Lymphocytes |  |  |  |  |
| low |  | 86 |  | 48.86364 |
| normal |  | 73 |  | 41.47727 |
| high |  | 17 |  | 9.659091 |
| C Reactive Protein(CRP) Levels |  |  |  |  |
| normal |  | 60 |  | 37.97468 |
| high |  | 98 |  | 62.02532 |
| CT Scan Results |  |  |  |  |
| pos |  | 124 |  | 89.20863 |
| neg |  | 15 |  | 10.79137 |
| RT-PCR Results |  |  |  |  |
| pos |  | 100 |  | 96.15385 |
| neg |  | 4 |  | 3.846154 |
| X-ray Result |  |  |  |  |
| pos |  | 35 |  | 74.46809 |
| neg |  | 12 |  | 25.53191 |
| GGO |  |  |  |  |
| Yes |  | 92 |  | 96.84211 |
| No |  | 3 |  | 3.157895 |
| Diarrhea |  |  |  |  |
| Yes |  | 30 |  | 45.45455 |
| No |  | 36 |  | 54.54545 |
| Fever |  |  |  |  |
| Yes |  | 261 |  | 91.25874 |
| No |  | 25 |  | 8.741259 |
| Coughing |  |  |  |  |
| Yes |  | 164 |  | 82.82828 |
| No |  | 34 |  | 17.17172 |
| Shortness Of Breath |  |  |  |  |
| Yes |  | 45 |  | 60 |
| No |  | 30 |  | 40 |
| Sore Throat |  |  |  |  |
| Yes |  | 37 |  | 60.65574 |
| No |  | 24 |  | 39.34426 |
| Nausea/Vomiting |  |  |  |  |
| Yes |  | 18 |  | 52.94118 |
| No |  | 16 |  | 47.05882 |
| Pregnant |  |  |  |  |
| Yes |  | 43 |  | 66.15385 |
| No |  | 22 |  | 33.84615 |
| Fatigue |  |  |  |  |
| Yes |  | 8 |  | 61.53846 |
| No |  | 5 |  | 38.46154 |

### 2.2 Relationship between pairs of clinical variables

We performed correlation between all possible pairs of clinical variables to uncover potentially important associations (Table S1). If both variables are continuous, the Spearman correlation test is applied (*p*<0.05). Among 143 Spearman correlation tests, 27 show significant correlation, with 9 that involve age, corroborating reports that age plays a critical role in the development of COVID-19[9]. We observed C reactive protein (CRP) levels and serum platelets levels to be the variables with the strongest correlations with age (Figure 1A). CRP levels, an indicator of inflammation, are positively correlated with increasing age, while platelets levels are negatively correlated with increasing age. Other than age, we observed a negative correlation between the levels of CRP and lymphocytes levels and a positive correlation between CRP levels and neutrophil levels (Figure 1A). This result suggests that inflammation is most likely driven by neutrophils. The serum levels of white blood cells are also strongly correlated with neutrophils, further suggesting that white blood cell counts are heavily influenced by neutrophil levels (Figure 1A).

**Figure 1.**
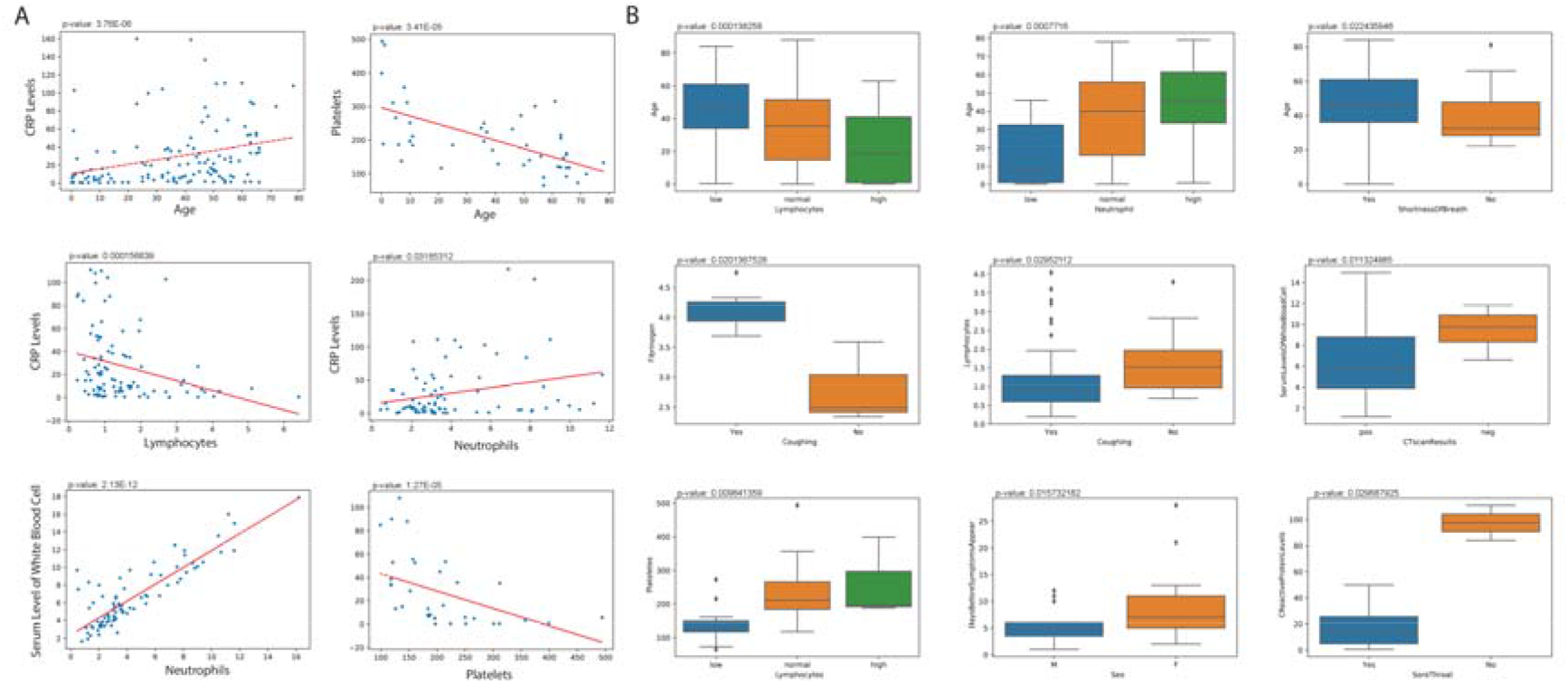
Select correlations with continuous clinical variables for COVID-19 patients. **A**. Correlations between two continuous variables (Spearman, *p*<0.05). **B**. Correlations between one continuous and one categorical variable (Kruskal-Wallis test, *p*<0.05).

For pairs of one continuous and one discrete variable, the Kruskal-Wallis test is applied (*p*<0.05). Among 319 Kruskal-Wallis comparisons, 36 correlations were significant. Some of the significant pairs overlapped with correlations between two continuous variables for variables that have both ordinal and continuous components. Such correlations are not displayed twice in Figure 1. We found again that age correlated significantly with multiple variables, including negative correlation with lymphocyte levels, positive correlation with neutrophil levels, and positive correlation with shortness of breath (Figure 1B). Other interesting associations were also discovered. Coughing was found to be correlated with increasing fibrinogen levels and decreasing lymphocyte levels. Those with lower levels of serum white blood cells (leukocytes) are more likely to report a positive CT scan result for pneumonia. Females may experience a greater number of days before symptoms appear. Finally, we found that sore throat decreases with increasing CRP levels (Figure 1B).

For pairs of two categorical variables, a two-tail chi-square test is applied. 42 out of 309 comparisons showed significant correlation, with few overlaps with former tests. Gender is involved in 6 of the significant correlation, indicating significant gender differences in COVID-19. Contingency tables of selected significant correlations are shown in Figure 2. Males were found to have higher lymphocyte and neutrophil levels than females (Figure 2A,B). Females were found to be more likely to have lower levels of serum white blood cells (Figure 2C).

**Figure 2.**
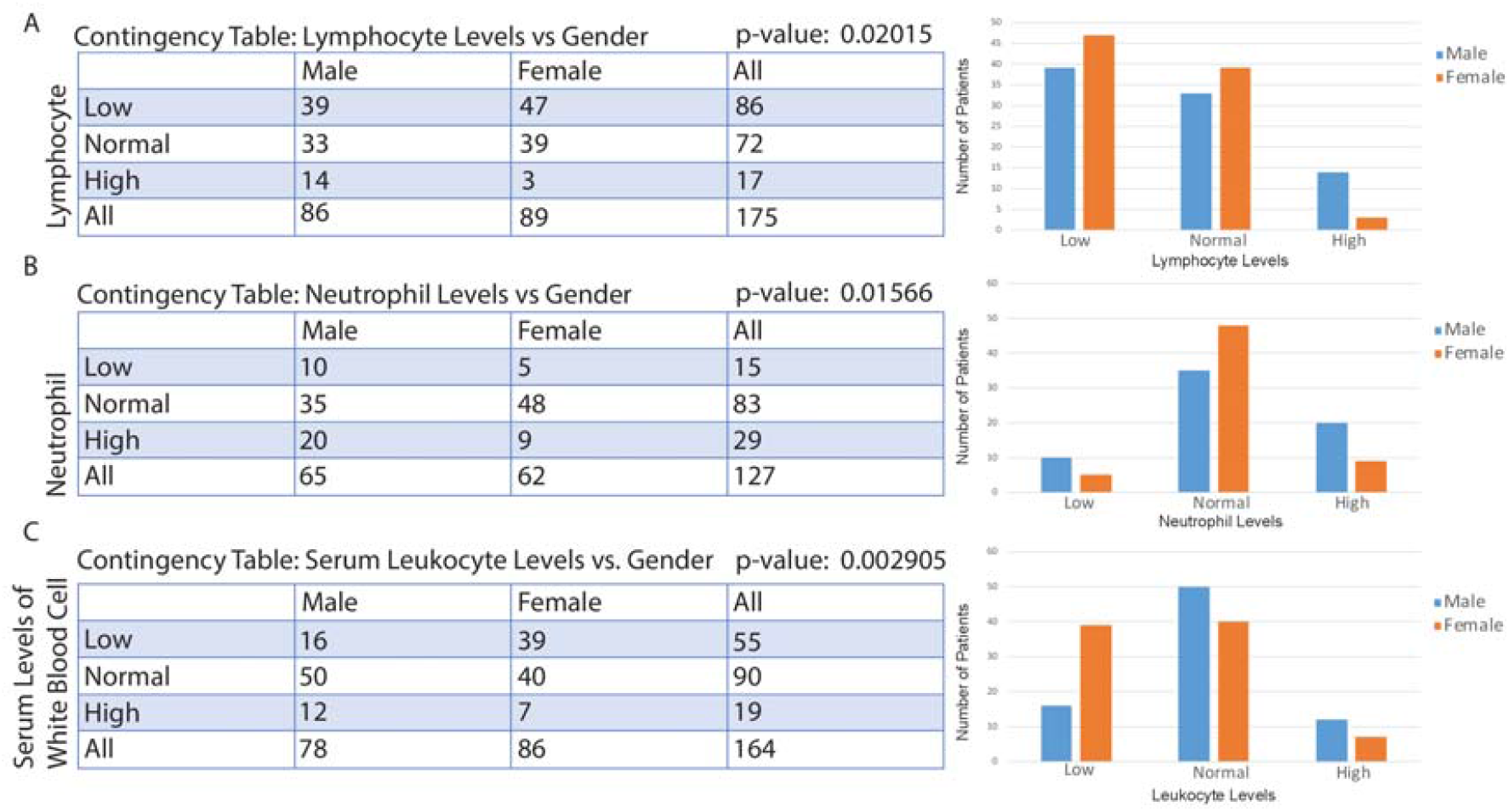
Correlations between gender and another categorical variable. **A**. Correlation between lymphocyte level categories and gender. **B**. Correlation between neutrophil level categories and gender. **C**. Correlation between serum leukocyte level categories and gender. A contingency table and a bar plot of the number of patients in each level are displayed for each correlation.

### 2.3 Clustering of patients into subcategories of COVID-19

We next aim to cluster COVID-19 patients based on clinical variables using machine learning. We chose the well-known SOM algorithm for clustering. SOM is a neural network that has a set of neurons organized on a 2D grid[10]. All neurons are connected to all input units (individual patients) by a weight vector. The weights are determined through iterative evaluations of a Gaussian neighborhood function, with the result of creating a 2D topology of neurons to model the similarity of input units (individual patients). The algorithm outputs a map that assigns each sample to one of the neurons on the 2D grid, with samples in the same neuron being the most similar to one another. Similarity of samples decrease with distance between neurons on the 2D map. Missing variables will be ignored from the SOM model when deriving a neural topology.

We generated square SOM neuron grids with side lengths 3 through 20 using the trainSOM function in the R package SOMbrero. The grids with side lengths 3, 4, 5, 7, and 9 all had topographic errors of 0 (Figure 3A). Of these, we chose the biggest grid (9×9=81 clusters) as our model. After the patients were assigned to neurons, an analysis of variance (ANOVA) test was performed to test which variables actively participate in the clustering. Of the 48 clinical variables we inputted, 27 were found to have very high significativity (*p*<0.001) (Table S2). We then reran the SOM using the 27 variables on a 9×9 grid. This grid is displayed on Figure 3B and has a final energy of 8.139248. The largest neuron has 39 patients, the second largest has 37, the third largest has 21, and the fourth largest has 20 (Figure 3C).

**Figure 3.**
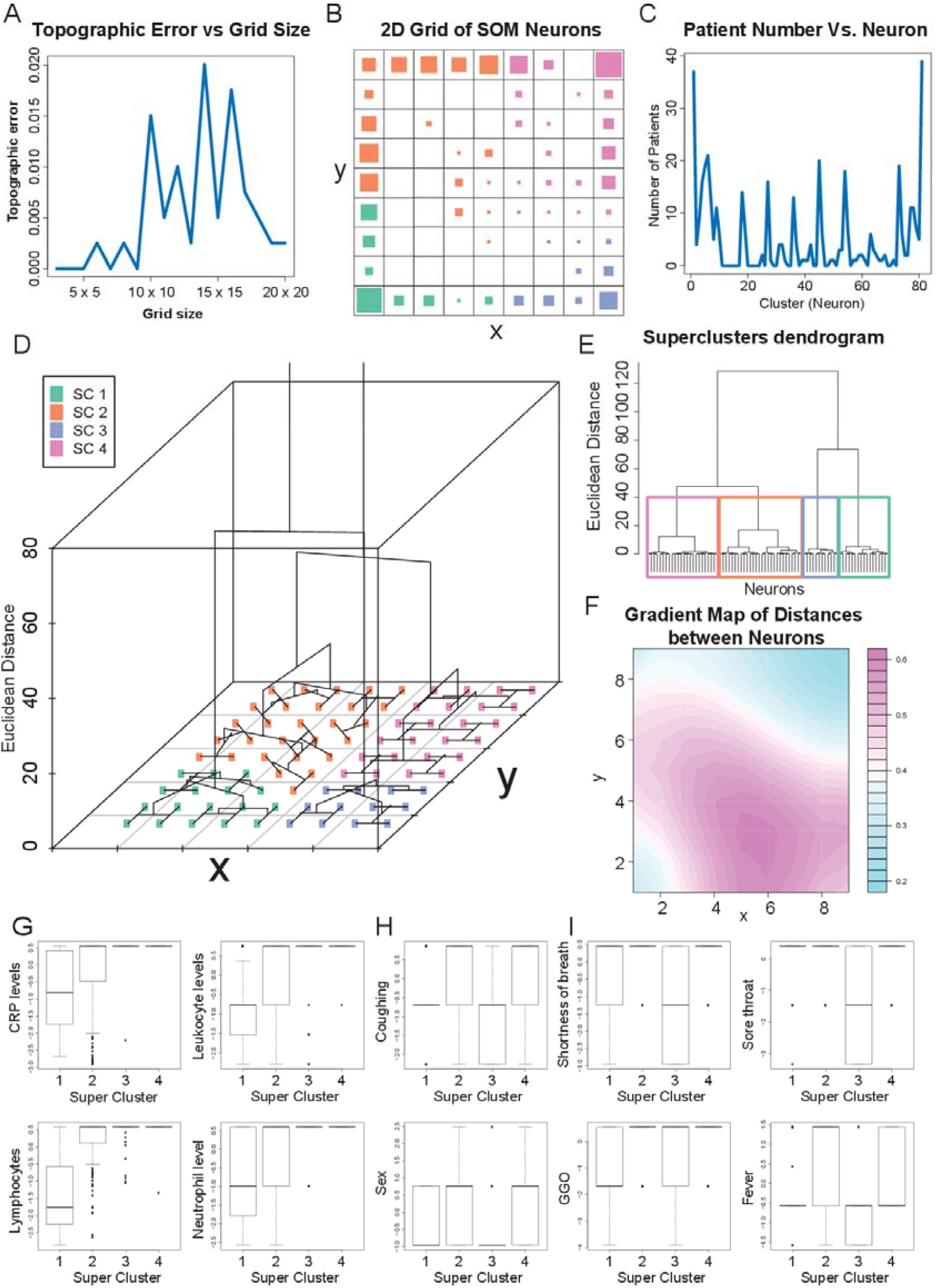
Summary of COVID-19 patient clustering using SOM. **A**. Plot of topographic error of the 2D SOM grid vs. size of the grid. **B**. 2D plot of SOM neurons after retaining only the most significant clinical variable for analysis. Each small grid represents a neuron, and the size of the square in each grid represents the number of patients associated with each neuron. The color code corresponds to superclusters presented in panel D. C. Plot of number of patients in each neuron. D. 3D dendrogram summarizing the neurons into superclusters. **E**. 2D dendrogram with the same information as the dendrogram in part D. In both dendrograms, the vertical axis represents the relative distance between clusters, which can be known between any two clusters by looking at the branch point where they diverge. **F**. Gradient map where light blue regions of the SOM depict higher similarity of neurons with each other. **G**. Boxplots of immune-associated clinical variables that differentiate superclusters. H. Boxplots in which superclusters 1 and 3 display similar trends.**I**. Boxplots in which only one supercluster has a median at a different value from the other three. All variables have been previously normalized. For binary variables, only three possible positions on the vertical axis is possible: the bottom one being no, the middle one being yes, and the top one being missing. For the gender (sex) variable, the bottom position is female, the middle is male, and the top one is missing.

### 2.4 Clinical characteristics of COVID-19 clusters

We then examined the defining features of patients assigned to the same neurons. We investigated four neurons associated with the largest number of patients and identified the four variables with the smallest nonzero standard deviations for each patient cluster. In the largest cluster, with 39 patients, the four smallest nonzero standard deviations were for the variables region of infection, sore throat, RT-PCR results, and coughing. In the second largest cluster, with 37 patients, the variables were baby death if pregnant, lymphocyte levels, fever, and coughing. In the next largest cluster, with 21 patients, the variables were sore throat, duration of illness in days, RT-PCR results, and coughing. In the fourth largest cluster, with 20 patients, the variables were sore throat, fever, coughing, and age.

We next used the function superClass to compute the relative Euclidean distances between the 81 patient clusters and form superclusters. The relative distances between the individual clusters are shown in Figure 3D-E. We divided the 81 clusters into 4 superclusters, which are represented in Figure 3B by the color of the squares. Supercluster 1 was formed with 24 neurons, supercluster 2 had 28 neurons, supercluster 3 had 12, and supercluster 4 had 17 neurons. Visualizing distances between neighboring neurons, we found that the distances are the smallest at corners of the grid, especially the upper right-hand corner (Figure 3F). This corner corresponds to supercluster 4, suggesting that patients within this cluster may be especially similar.

Next, we sought to determine the clinical features that effectively distinguish these superclusters. We performed Kruskal-Wallis testing on the values of the 27 variables across the four superclusters. 24 variables were significantly different between the superclusters (*p*<0.05) (Table S3). We discovered that the clinical variables exhibit 3 main types of correlations with the superclusters: continuous increase in value from cluster 1 to cluster 4 (Figure 3G), clusters 1 and 3 exhibiting the same distribution and clusters 2 and 4 exhibiting another distribution (Figure 3H), and 3 clusters exhibiting the same median (Figure 3I). From these analyses, we could infer that patients with low levels of CRP and serum immune cells likely define cluster 1. Cluster 1 patients are also predominantly female. Cluster 2 contains patients with slightly higher levels of CRP and serum immune cells than cluster 1. Compared to cluster 1 patients, fewer cluster 2 patient report coughing and fever. Cluster 2 patients are predominantly male. Cluster 3 contains patients with few reported symptoms, including less coughing, shortness of breath, fever, and sore throat. Cluster 3 is overwhelmingly female. Cluster 4 most likely contains patients not belonging to the other 3 clusters as it has few distinguishing features and high levels of missing data.

### 2.5 Creation of a diagnostic model for COVID-19 based on clinical variables

Because it can be difficult to distinguish influenza from COVID-19, we downloaded clinical data collected for influenza from a study by Cheng et. al. and from the Influenza Research Database[11, 12]. Machine learning was then used to perform a classification task to discriminate between influenza and COVID-19. For machine learning, we employed the algorithm Extreme Gradient Boosting (XGBoost) using Python. XGBoost is a novel, state-of-the-art machine learning algorithm that has been shown to outperform other more traditional algorithms in its accuracy and efficiency[13]. It can also take both continuous and discrete inputs and handle sparse data, in addition to having highly optimizable hyper-parameters[14].

The datasets from non-COVID patients and COVID-19 patients were merged and then split into training and testing patient sets, with 80% and 20% of the patients, respectively. Categorical variables were encoded as dummy variables. We then tuned the model using the Bayesian optimization method for hyperparameter search. We found the best hyperparameters to be gamma=0.0933, learning rate= 0.4068, max depth=6.558, and n_estimators=107.242.

### 2.6 Evaluation of XGBoost classification outcomes

From the ROC curve of prediction results, we obtained an AUC of 0.990 (Figure 4A). However, because there is an imbalance of class in our input (i.e. we have significantly more influenza patients than COVID-19 patients), the precision recall (PR) curve may be better able to present our model’s results. ROC curves could be significantly influenced by skewing the distribution of classes in classification, while PR curves would not be impacted by this action. We observed a slightly lower AUC of 0.977 in our PR curve and computed the F1 score to be 0.929 (Figure 4B), which suggest that our model is still highly accurate even when class imbalances are taken into account. The prediction result from XGBoost’s predict function was used to plot a confusion matrix (Figure 4C). From the confusion matrix, we calculated a sensitivity of 92.5% and a specificity of 97.9%. We found the most important features in our prediction model to be age, CT scan result, temperature, lymphocyte levels, fever, and coughing, in order of decreasing importance (Figure 4D). We also provided a 6-level decision tree sample of our XGBoost model (Figure 4E), which is not a representation of our full model.

**Figure 4.**
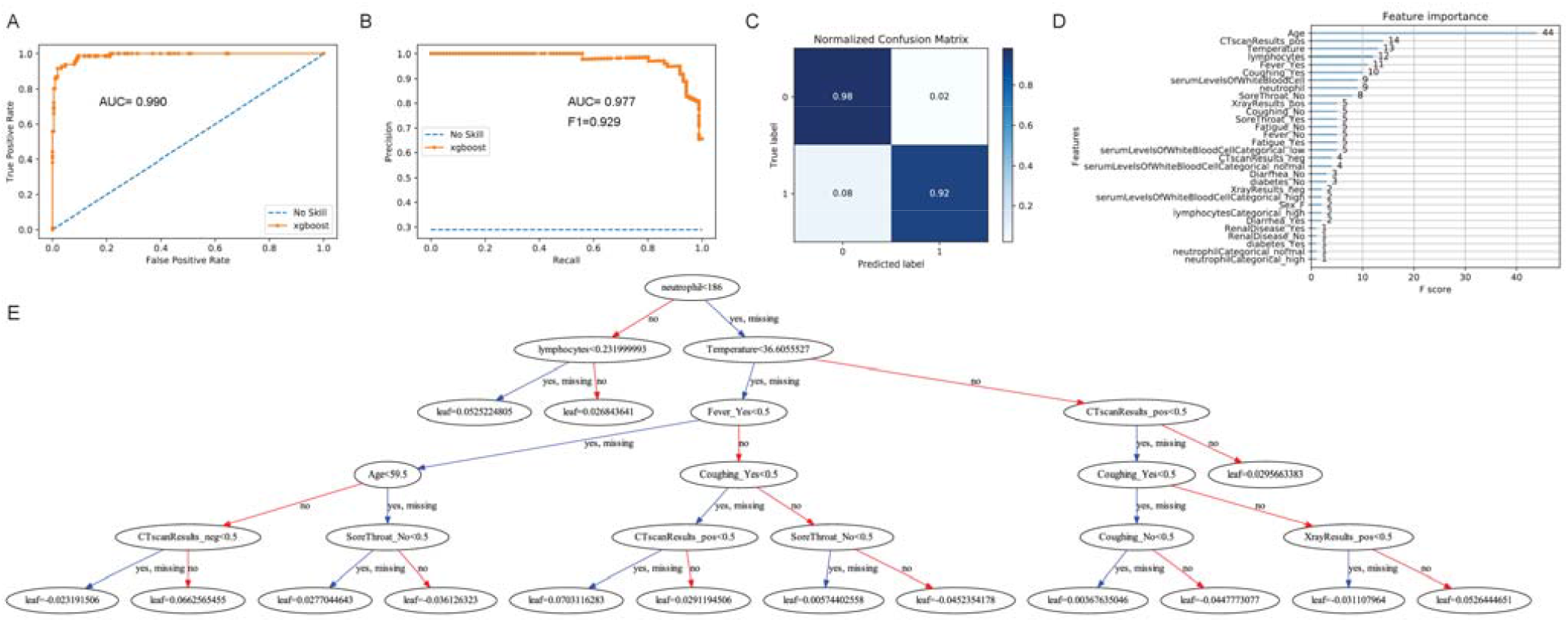
Summary of XGBoost classification of COVID-19 and influenza patients. **A**. ROC curve of prediction. **B**. Precision recall curve of prediction. **C**. Confusion matrix of prediction. **D**. Variables most important for classification, listed by decreasing order of importance. **E**. 6-level sample model of SOM decision tree construction.

## 3. Discussion

As the recent pandemic of COVID-19 unfolds across the world, the inability of countries to test their citizens is heavily impacting their healthcare system’s ability to fight the epidemic. Testing is necessary for the identification and quarantine of COVID-19 patients. However, the multi-step process required for the conventional SARS-Cov-2 test, via quantitative polymerase chain reaction (qPCR), is creating difficulties for countries to test large numbers of suspected patients[15]. Testing begins with a healthcare worker taking a swab from the patient. The swab is sent to a laboratory, and viral RNA is extracted from the sample and reverse transcribed into DNA. The DNA is tagged with a fluorescent dye and then amplified using a qPCR machine. If a high level of fluorescence is observed compared to control, the sample is positive with SARS-Cov-2. Each step of the testing process is susceptible to severe shortages[16].

In this study, we aim to mine published clinical data of COVID-19 patients to generate a new diagnostic framework. We hypothesize that novel or complex associations between clinical variables could be exploited for diagnosis with the aid of machine learning. Not only may underlying relationships between clinical variables in COVID-19 be useful for the development of a computational diagnostic test based on signs, symptoms, and laboratory results, these correlations can also yield critical insights into the biological mechanisms of COVID-19 transmission and infection.

Using correlational tests, we corroborated previous findings and expected results for COVID-19 patients but also uncovered novel relationships between clinical variables. We found that age is correlated with CRP level, an indicator of inflammation, and decreased platelet levels. It is known that as age increases, the proinflammatory response becomes stronger, leading to increasing CRP and decreasing platelet levels[17]. However, we found surprising correlations with gender, including higher serum neutrophil and leukocyte levels in males compared to females. According to the National Health and Nutrition Examination Survey, with data from over 5,600 individual, few differences exist between male and females in the serum levels of these cells[18]. Another study with 200 samples found that neutrophils are generally higher in women[19]. Correlations with gender observed here may offer a piece of the explanation for why men infected with COVID-19 seem to experience poorer prognosis, one of the important outstanding questions of COVID-19[20].

We also classified COVID-19 patients into different clusters using the SOM machine learning algorithm. Two of the clusters are defined by low vs. high levels of immunological parameters, including immune cell counts and CRP levels. A third cluster is defined by a tendency for less reported symptoms, including sore throat, fever, and shortness of breath, and is predominantly female.

Finally, using the machine learning algorithm XGBoost, we constructed a computational model that successfully classified influenza patients from COVID-19 patients with high sensitivity and specificity. We believe that our model demonstrated the feasibility of using data mining and machine learning to inform diagnostic decisions for COVID-19. Such a model could be extremely useful for more effective identification of COVID-19 cases and hotspots, which could allow health officials to act before testing shortages could be addressed.

Despite promising results, several limitations exist for our study, all of which stem from the lack of large-scale clinical data. First, our sample size is severely limited because most clinical reports published do not publish individual-level patient data. Second, data on influenza signs and symptoms are equally inaccessible. We were only able to locate data for patients with H1N1 influenza A, which is not one of the active strains in the current influenza season. Third, many of our data sources are case studies that focused on specific cohorts of COVID-19 patients. This increases the chance of us capturing a patient population that is not representative of the general population, although this is an inherent risk of sampling. We anticipate that as more data are made openly available in the weeks and months to come, we will be able to build a more robust computational model. Therefore, we intend to provide the model we constructed as a computational framework for computation-aided diagnosis of COVID-19 data rather than a ready-to-use model. We also encourage researchers around the world to release de-identified patient data to aid in data mining and machine learning efforts against COVID-19.

## 4. Materials and Methods

### 4,1 Literature search and inclusion criteria for studies

Patient clinical data were manually curated from a PubMed search with the keyword “COVID-19.” A total of 1,439 publications, dating from January 17, 2020 to March 23, 2020, were reviewed. All publications with no primary clinical data, including reviews, meta-analyses, and editorials, were excluded from our analysis. After manual review, we found 151 studies with individual-level data, encompassing data from 413 patients. All individual patient data with 2 more clinical variables reported per patient were included. Clinical variables sought for included demographics, signs and symptoms, laboratory test results, imaging results, and COVID-19 diagnosis. No formal review protocol was used. No bias was assessed within or across studies because we did not include any clinical trials or case-control studies.

For our machine learning classification task to discriminate COVID-19 patients from influenza patients, we used clinical variables for 21 influenza patients from a study by Cheng et. al. and 1050 patients from the Influenza Research Database[11, 12]. Only H1N1 Influenza A virus cases were included because of difficulties locating data from other strains.

### 4.2 Correlational tests between pairs of clinical variables

We sought to uncover correlations that could yield critical insights into the clinical characteristics of COVID-19 by correlating every variable to each other. For two continuous variables, the Spearman correlation test was applied. For one continuous variable and one categorical variable, the Kruskal-Wallis test was applied. For two different categorical variables, the chi-squared test will be applied. All statistical tests were considered significant if the p-value is 0.05 or below.

### 4.3 Machine learning for classification of COVID-19 patients into subtypes

A self-organizing map (SOM) is an artificial neural network that constructs a two-dimensional, discretized depiction (map) of the training set. We used the SOM algorithm to cluster our patients based on similar patterns of clinical variables. The SOMbrero package in R was used[21]. Because clustering of neurons are performed using Euclidean Distance, we first standardized each clinical variable to ensure that they are equally weighted.

The trainSOM function was used to implement numeric SOM on our data set, which is inputted as an N × P matrix, with N=398 patients and P=48 variables. From this, we selected 27 clinical variables with very high significativity (*p*<0.001) after running an ANOVA test across all neurons and ran another iteration of trainSOM with these variables. We generated SOMs from the 3×3 neuron grid to 20×20 neuron grid and selected the 9×9 SOM with 81 neurons as our final model based on minimal topographic error. We then aggregated the neurons into super-clusters using the superClass method in SOMbrero.

### 4.4 Preprocessing of data for machine learning classification

Data were preprocessed by combining data from COVID-19 cases and influenza cases into a single matrix, followed by removal of any clinical variables that were not present in both the COVID-19 dataset and the influenza dataset. 19 clinical variables were included as machine learning input. The variables include age, sex, serum levels of neutrophil (continuous and ordinal), serum levels of leukocytes (continuous and ordinal), serum levels of lymphocytes (continuous and ordinal), result of CT scans, result of chest X-rays, reported symptoms (diarrhea, fever, coughing, sore throat, nausea, and fatigue), body temperature, and underlying risk factors (renal diseases and diabetes). Categorical data were converted to dummy variables using the get dummies function in Pandas because non-numerical data are not allowed in our machine learning algorithm.

### 4.5 Performing XGBoost classification

The eXtreme Gradient Boosting algorithm (XGBoost), an ensemble machine learning method widely known for its superior performance over other machine learning methods, was selected for our study[13]. We first split our data into 80% training dataset and 20% testing dataset. 5-fold cross-validation was then performed, with 70 boosting rounds (iterations), and fed into a Bayesian optimization function for calculation of the best hyperparameters for XGBoost. The hyperparameters tuned included max depth, gamma, learning rate, and n estimators. Bayesian optimization was performed with an initial 8 steps of random exploration followed by 5 iterations. The expected improvement acquisition function was used.

### 4.6 Evaluation of XGBoost results

XGBoost results were evaluated by plotting a receiver operating characteristic (ROC) curve and a precision recall (PR) curve. The area under the curve (AUC) was also calculated for both curves.

## Data Availability

The datasets during and/or analysed during the current study available from the corresponding author on reasonable request.

## Declarations

### Ethics approval and consent to participate

Not applicable.

### Consent for publication

Not applicable.

### Author Contributions

Conceptualization, W.M.O.; methodology, W.M.O. and W.T.L.; software, W.T.L. J.M., and N.S.; validation, W.T.L., J.M., and N.S.; formal analysis, W.T.L., J.M., and N.S.; investigation, W.T.L, J.M., N.S., G.C., J.C., J.C.T., L.A., C.O.H., J.X., L.M.W., T.Z., A.L., A.G., and T.K.H.; resources, W.M.O.; data curation, W.T.L, J.M., N.S., G.C., J.C., J.C.T., L.A., C.O.H., J.X., L.M.W., T.Z., A.L., A.G., and T.K.H.; writing—original draft preparation, W.T.L. and N.S.; writing—review and editing, W.M.O., E.Y.C., M.R.R., S.Z.K., and M.A.Y.; visualization, W.T.L., J.M., and N.S.; supervision, W.M.O.; project administration, W.M.O.; funding acquisition, E.Y.C. All authors have read and agreed to the published version of the manuscript.

## Acknowledgments

Not applicable.

## Abbreviations

CRP: C-reactive Protein
ANOVA: Analysis of Vatriance
SOM: Self-organizing map
XGBoost: Extreme Gradient Boosting
ROC: Receiver Operating Characteristic
AUC: Area Under the Curve
PR: Precision-Recall

